# *“WE CAN ALL CONTRIBUTE IN OUR OWN WAY”* : KNOWLEDGE MOBILIZATION TOOLS TO PROMOTE BEST PRACTICES IN UNIVERSAL ACCESSIBILITY

**DOI:** 10.1101/2024.09.17.24313810

**Authors:** Maëlle Corcuff, François Routhier, Marie-Eve Lamontagne

## Abstract

**Background:** Cities aim to enhance urban accessibility following the adoption of the United Nations’ Convention on the Rights of Persons with Disabilities. However, implementation faces challenges due to complex municipal legislation, lack of awareness, and organizational obstacles. Engaging stakeholders and empowering municipal employees through knowledge mobilization is crucial, as shown in a Quebec City’s partnership research process.

**Aim:** To report the implementation strategy as implemented, explore the perception of the employees about the format and feasibility of the implementation strategy and explore the induced changes of knowledge mobilization tools on the implementation determinants of universal accessibility measures for municipal employees.

**Methods:** The study used a multi-method design, involving interviews and a questionnaire with the project steering committee, made up of city employees and the research team. Three 30-minute participatory workshops were conducted for culture, communications, and public consultation administrative units.

**Results:** Participants appreciated the workshop format and video content, suggesting minor improvements for broader implementation. The tools effectively increased engagement in implementing universal accessibility measures, proving valuable for raising awareness.

**Discussion and Conclusion:** The study demonstrates the advantages of a collaborative approach in developing knowledge mobilization tools, enhancing municipal personnel’s capacity for universal accessibility measures, and highlighting the need for adaptable strategies.

Contributions to the litterature

- Knowledge mobilization tools created in partnership with knowledge users encourage buy-in and a positive view of the tools.
- An interactive implementation strategy actively involving knowledge users promotes awareness and behavior change
- Municipal organizations’ context being complex, the implementation strategy must be adapted to each group of people and their reality to facilitate the implementation and adoption of the tools.
- The combined use of a theoretical framework and a participatory approach provides a guideline for the development of tools and implementation, while adapting to the specific context.

## INTRODUCTION

Cities increasingly recognize the importance of implementing policies aimed at enhancing the universal accessibility of their urban environments, encompassing services, infrastructures, and communications [1–4]. Since the adoption in 2010 of the United Nations’ (UN) Convention on the Rights of Persons with Disabilities [1], the universal accessibility of urban spaces has emerged as a significant public concern, particularly with demographic shifts and the rising population of individuals potentially living with disabilities [5, 6].

The positive impacts of universally accessible environments reverberate through many aspects of community life, creating a more equitable, dynamic, and resilient society [7]. Investment in universally accessible environments is a driver of social, economic, and cultural progress and a collective mobilization towards an inclusive society [8]. Consequently, it becomes essential to design environments that can be used by all, fostering engagement, and strengthening the ability of all individuals, regardless of their abilities, to fully participate in their community [9]. Urban environments are major hubs for job creation, transportation networks, implementation of innovations and provision of services to populations [10, 11]. Municipal governments must therefore adopt policies to facilitate the inclusion of all citizens. In order to implement these policies, municipal employees, coming from different backgrounds, must develop strategies to adopt best practices to set up accessible environments [9, 12].

However, the implementation of universal accessibility measures planned in municipal legislations faces many challenges [13, 14]. The lack of knowledge, education and awareness about the various issues involved in creating accessible and inclusive environments experimented by decision-makers and municipal employees might complicates the implementation of planned measures [15]. As a result, measures to improve the environment tend to focus less on user needs, capabilities and pathways, and more on sustainability, form, function and budget [14]. For example, urban designers tend to associate universal accessibility with a form of restriction to their creativity [16]. They also rely on their own interpretation of the experiences of people concerned by lack of accessibility [17]. There is a significant lack of evidence-based tools, methods, and processes to help the stakeholders involved gather information and assess and improve the accessibility of their infrastructures and services [18]. In addition, the organizational complexity of municipal organizations is a major obstacle to the implementation of universal accessibility measures [19]. Indeed, the important number of administrative units, the political priorities and the different hierarchical layers in the organization can slow down the adoption of new and better universal accessibility practices [20].

In light of the above, it is important to promote the efficient implementation of universal accessibility policies by mobilizing the stakeholders involved [17], to empower municipal employees in what they can do in terms of universal accessibility [21]. Research has shown that knowledge mobilization is an effective catalyst for the implementation of evidence-based services and policies in various contexts [9, 15, 22, 23]. Authors also demonstrate that the creation of knowledge mobilization tools within municipalities and about universal accessibility practices has positive repercussions on the social, economic and health dimensions of populations [2, 6, 15, 24] that contributes to alleviate the challenges faced municipalities. The development and creation of context-specific knowledge mobilization tools would therefore be a facilitator for the implementation of the universal accessibility measures set out in policies.

This study took place in a city in the province of Quebec, with a population of around 550,000 citizens and a metropolitan community of about 840 000 people. This municipal organization has 5,000 full-time employees working as managers, civil servants, professionals, technicians, workers, or seasonal workers in 33 administrative units. This city engaged in a partnership research process to overcome obstacles encountered by its employees toward universal accessibility implementation by co-designing knowledge mobilization tools. The co-design process used with partners facilitates stakeholder buy-in and involvement in the research process [25] and enable and a better match between the creation of suitable tools and the response to existing needs [26, 27]. The co-design process resulted in the production of an optimal implementation strategies within workshops, presenting three short videos about universal accessibility within the city as the knowledge mobilization tools [27] to promote awareness and understanding of universal accessibility.

This article aims to 1) report the implementation strategy as implemented, 2) explore the perception of the employees about the format and feasibility of the implementation strategy and 3) explore the induced changes of knowledge mobilization tools on the implementation determinants of universal accessibility measures for municipal employees.

## METHODS

We used a multi-method design [28] with semi-structured interviews and an electronic questionnaire. We also used a partnership approach with the project steering committee, made up of city employees and the research team along the study. The results reported in this study are based on implementation and evaluation in three administrative units. This served as a pre-test of the implementation strategy with a view to mass deployment of knowledge mobilization tools across all administrative units once the pre-test will be carried out and evaluated.

### 1. Implementation strategy of the tools

To obtain an adapted, coherent, and solid implementation strategy, the steering committee chose to use a logic model to structure the strategy. As recommended by Kellogg Foundation [29], they first set out the objectives of the implementation strategy and then identified the activities required to achieve each objective. Figure 1 shows the implementation strategy embedded in the logic model developed by the steering committee.

**Figure 1.**
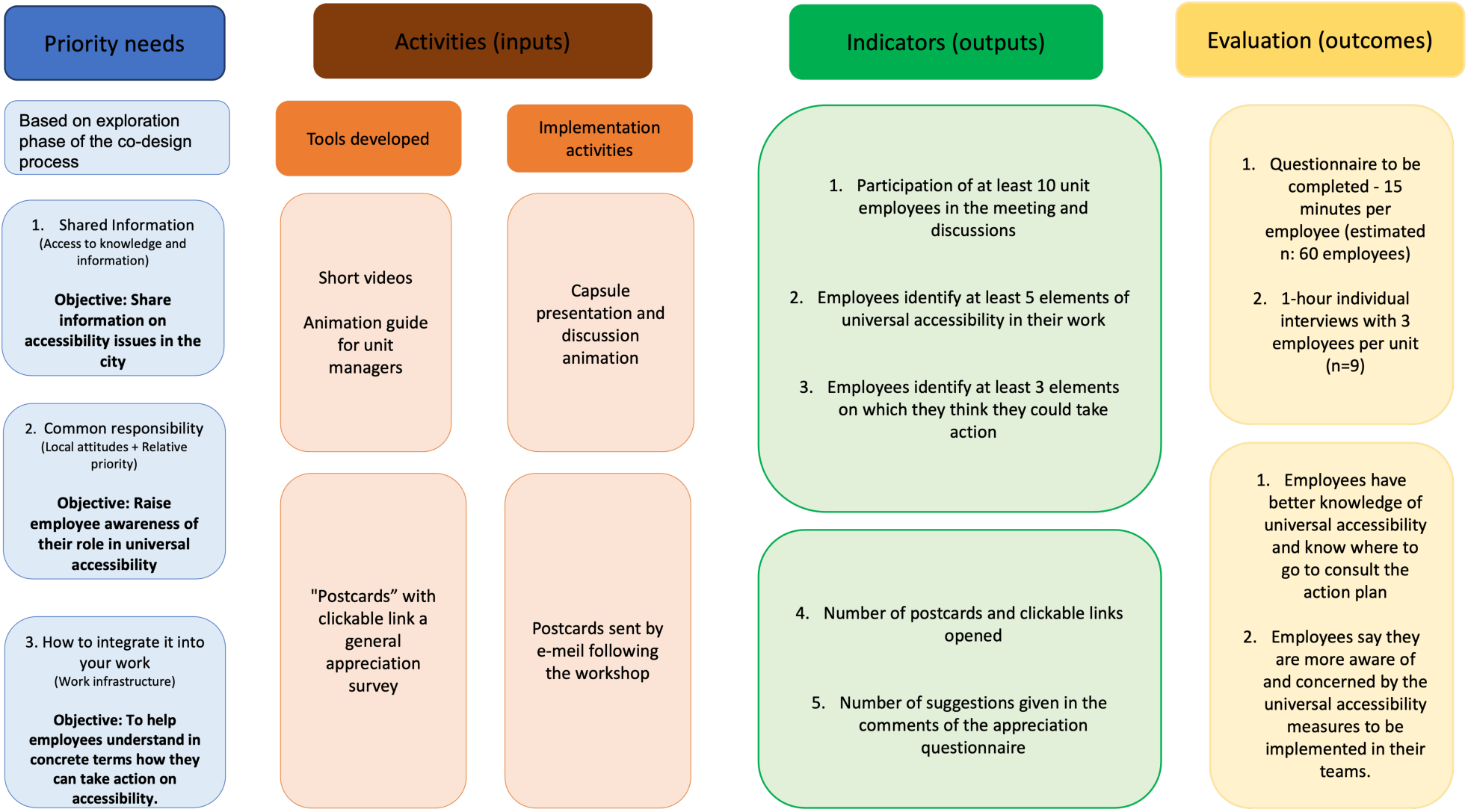
Logic model of implementation strategy.

#### Participants

The selection of the three test administrative units was based on the composition of the steering committee team, driven by feasibility constraints regarding deadlines. Workshops were conducted with the culture, communications, and public consultation units, as these groups are significantly impacted by universal accessibility concerns and regularly engage with citizens. The researcher (MC) attended all workshops in an observational posture. Discussions during the workshops were recorded and transcribed for inductive content analysis using *NVivo 14* [30]. Additionally, brief meetings with workshop facilitators were held after each session and at the conclusion of the series to collect their feedback in a logbook.

#### Activities and output

In consideration of the busy schedules of the municipal administrative units and to enhance engagement, three 30-minute participatory implementation workshops were scheduled for each team. Some administrative units participated in the pre-test of these workshops. Workshops are recognized as moderately effective dissemination strategies in implementation science [31, 32]. The participatory workshops unfolded in two segments. First, a brief overview and project context were provided, followed by the presentation of short videos to the team. Discussions and exchanges on the videos were then led by the team’s universal accessibility manager, using an animation guide developed by the steering committee. At this stage, the first author (MC) recorded the discussions in a logbook. Workshops could be held in person or virtually, according to individual preferences. Following the workshops, the first author (MC) met the workshop facilitators via the Microsoft TEAMS platform and collected their feedback.

### 2. Evaluation procedures

First, a questionnaire based on the Consolidated Framework for Implementation Research (CFIR) [33], was distributed to all participants who attended the three workshops. This questionnaire aimed to explore the effect of the workshops and knowledge mobilization tools on participants’ knowledge, beliefs, perceptions, and skills regarding the implementation of universal accessibility practices. Responses were collected using a Likert scale [34, 35].

Then, the first author (MC) also conducted semi-structured interviews, from volunteers who have expressed an interest in participating by e-mail to the researcher. The interviews lasted approximately one hour and aimed to explore the perception of the employees about the format and feasibility of the implementation strategy. Participation in these interviews required only attendance at the three workshops. The interview guide contained knowledge transfer questions on format and content and was based on the CFIR. Participants provided informed consent.

### 3. Analysis

Logbook data from workshop discussions and exchanges with facilitators were examined through content analysis. Virtual interviews, conducted via the Microsoft TEAMS platform, were recorded, transcribed, and analyzed using *NVivo 14* [30] software through inductive content analysis. Questionnaire data underwent descriptive statistical analysis.

## RESULTS

### 1. Implementation strategy

The nine workshops were conducted virtually, except for one of the third session with the public consultation team, which was held in person. These workshops lasted in average 25 minutes, ranging from 16 to 38 minutes, including discussions, spanning a three-month period from November 2023 to January 2024. Participation varied across administrative units, with 31 members from the communication unit, 12 from the culture unit, and 18 from the public consultation unit. In total, 61 employees participated in the workshops. The Communications and Public Consultations administrative units met every two weeks, except for the third session, which occurred a month after the second due to the Christmas break. The Culture team’s workshops occurred within a one-month timeframe.

#### Workshop discussions

##### 1st workshop

During the first workshop, participants acknowledged that while they had prior notions of universal accessibility, the video’s concrete examples expanded their understanding, revealing elements of accessibility they hadn’t consciously noticed before. It also highlighted that universal accessibility impacts everyone, not just individuals living with disabilities. Employees perceived the video as a valuable awareness tool, suggesting its potential use to showcase the city’s achievements to citizens. They emphasized the importance of the city acknowledging its accomplishments while considering future improvements. Additionally, discussions sparked ideas for additional tools, such as checklists, accessibility indicators, and a database of available resources, to support employees further.

##### 2nd workshop

Most participants admitted they were unaware of the plan’s specifics or its relevance to their team before the workshop. They learned about existing services and expressed a desire for increased awareness among both employees and citizens. They emphasized the importance of familiarizing themselves with these services to better assist citizens and discussed the utilization of available tools, such as magnetic loops for the hearing-impaired, and how to promote these services within partner organizations. Participants also shared existing initiatives they had implemented and noted the video helped clarify appropriate terminology and underscored the importance of collaborative efforts in creating fully accessible pathways. They acknowledged that while ensuring accessibility requires initial effort, it will eventually become second nature, highlighting the necessity of proactive integration efforts. One participant stressed, "*Accessibility is the key to inclusion, and it’s essential that our administration takes the time to do it*."

##### 3rd workshop

This vignette was highly valued by participants, who noted they learned much about their colleagues’ accomplishments, previously unknown to them. They were surprised to discover the extent of universal accessibility efforts across various municipal administrative units. They appreciated the authenticity of real employees sharing their actions, finding it more impactful than reading about them in the action plan. Witnessing tangible achievements emphasized the importance of adopting universal accessibility practices and their positive impact on the environment and citizens. Participants stressed the need for widespread awareness and integration of universal accessibility principles into daily practices. While acknowledging the city’s progress, they recognized the ongoing need for improvement. One participant expressed *"I’m embarrassed because my activities aren’t always accessible, and I don’t have a plan B*" when comparing with colleagues in the video. However, one participant added that *"little things can make a big difference, more than you think*".

###### Facilitators’ feedback

The workshops moderators identified two main factors that could limit the depth of discussions and participant engagement. Firstly, larger groups, such as the 31-member communications team, tended to have fewer exchanges, and it was challenging to encourage their involvement despite follow-up questions. Additionally, conducting workshops virtually sometimes led to lower engagement levels compared to face-to-face sessions. The moderator of the public consultation team observed a notable increase in participation during the last in-person workshop compared to the preceding virtual sessions. Employees were more actively engaged in discussing universal accessibility issues, which could be partly attributed to familiarity with the content by the third session. The timing of the third workshop, after the Christmas break for two administrative units (public consultation and communications), necessitated a recap of previous videos and discussions to re-engage participants.

The facilitators emphasized the importance of tailoring workshop leadership to the attendees’ characteristics, such as their level of commitment, comfort with public speaking, existing knowledge of universal accessibility, and their interaction with citizens. They suggested incorporating ice-breaker activities, like "in the shoes of…", especially when workshops are part of broader team meetings covering multiple topics. Additionally, they stressed the need to clearly communicate the objectives of the workshops and videos from the outset and to reiterate them as necessary throughout the sessions.

### 2. Evaluation

#### 2.1. Questionnaire

A total of 44 employees answered the questionnaire, representing 72% of the employees who participated in the implementation strategy. 30 out of the 44 questionnaires were entirely completed. Among the respondents, 21% had participated in the initial survey before the presentation of the short videos, and 11% had taken part in focus group discussions aimed at better understanding their needs. Table 1 shows the job characteristics data of the participants who answered the questionnaire. Six participants did not complete this section of the questionnaire.

**Table 1.** Job characteristics of participants.

| Characteristics | All participants (n=44) |
| --- | --- |
| Time on the job |  |
| Less than 6 months | 1 (2.3%) |
| 6 months to 1 year | 5 (11.4%) |
| 1 to 5 years | 18 (40.9%) |
| 5 to 10 years | 2 (4.5%) |
| More than 10 years | 12 (27.3%) |
| No answer | 6 (13.6%) |
| Job category |  |
| Professional | 28 (63.6%) |
| Civil servant | 9 (20.5%) |
| Executive | 1 (2.3%) |
| No answer | 6 (13.6%) |
| Administrative unit |  |
| Public consultation | 8 (18.2%) |
| Communication | 19 (43.2%) |
| Culture | 11 (25.0%) |
| No answer | 6 (13.6%) |

**Table 1. Job characteristics of participants**
| **Characteristics** | **All participants**<br>**(n=44)** |
| --- | --- |
| Time on the job |  |
| Less than 6 months | 1 (2.3%) |
| 6 months to 1 year | 5 (11.4%) |
| 1 to 5 years | 18 (40.9%) |
| 5 to 10 years | 2 (4.5%) |
| More than 10 years | 12 (27.3%) |
| No answer | 6 (13.6%) |
| Job category |  |
| Professional | 28 (63.6%) |
| Civil servant | 9 (20.5%) |
| Executive | 1 (2.3%) |
| No answer | 6 (13.6%) |
| Administrative unit |  |
| Public consultation | 8 (18.2%) |
| Communication | 19 (43.2%) |
| Culture | 11 (25.0%) |
| No answer | 6 (13.6%) |

Among the respondents, 58% indicated an improved understanding of universal accessibility following the workshops, while 21% reported no change due to existing knowledge. Regarding the action plan for universal accessibility, 34% could explain and locate it, while 30% could explain but not locate it. Post-workshop, 43% incorporated universal accessibility measures based on needs, with 25% integrating them daily. Only one person expressed no need to incorporate universal accessibility into their work.

The questionnaire results also revealed, across various domains of the CFIR framework [33], the impact of integrating the short videos into workshops, as illustrate in the following figures. Figure 2 outlines the prevalence of participant agreement (shown in green) with different items related to the innovation and individual characteristics constructs of CFIR. Employees generally agreed with most items within these constructs. The complexity of the intervention was assessed in reverse. 55% of employees indicated that the workshops and short videos were not very complex to implement. Additionally, nearly half of the respondents did not provide feedback on the costs of the innovation, which is expected as it may not be relevant to their role.

**Figure 2.**
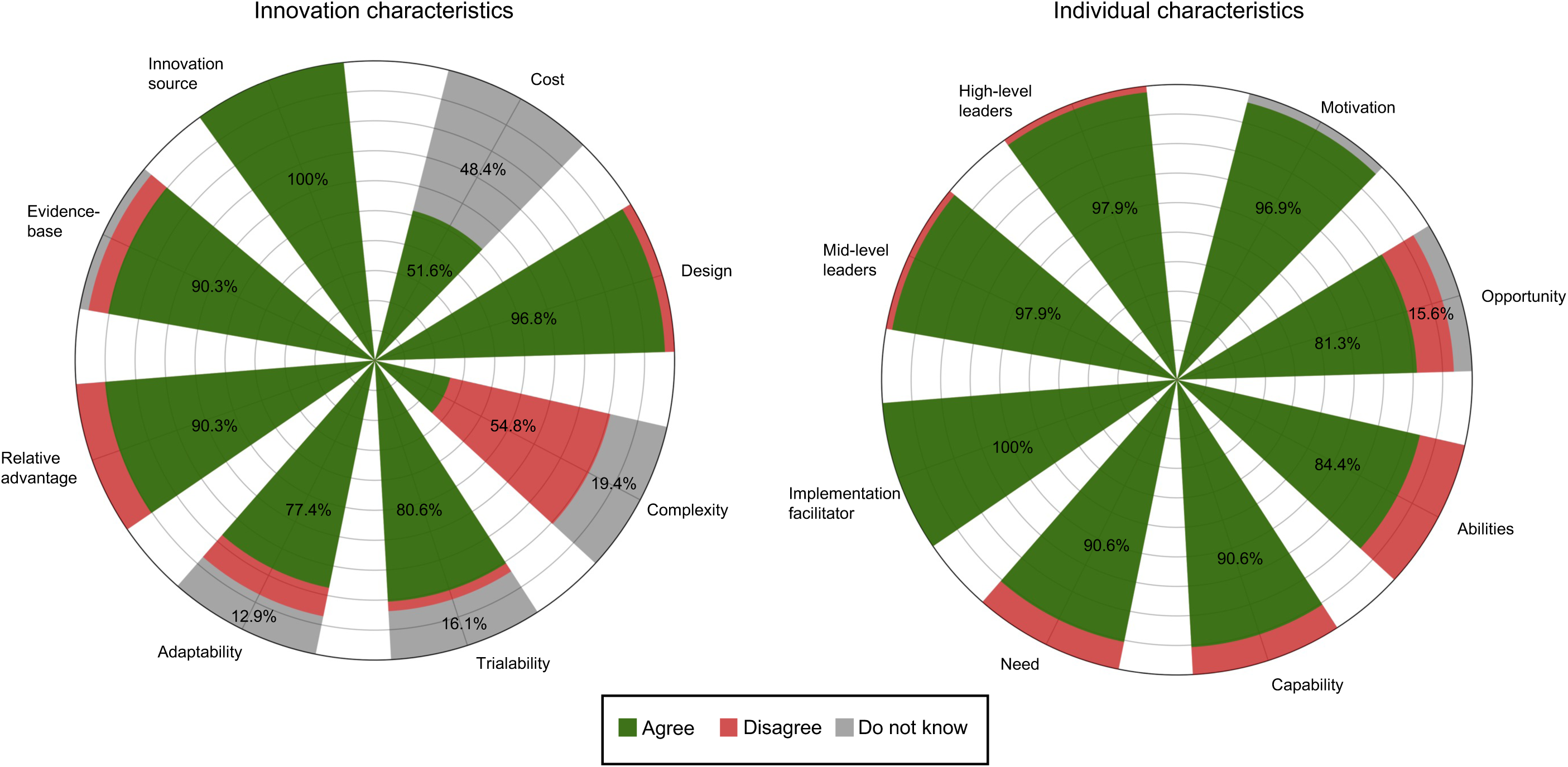
Agreement of participants with innovation and individual characteristics items.

Figure 3 depicts the facilitators and barriers to implementation as reported by employees regarding the internal and external context constructs of the organization following the workshops and short video presentations. It is evident that most constructs are facilitators after assisting to the workshops. Within the internal context, this is particularly notable for resources available to support implementation and the structural characteristics of the organization, such as the physical work environment. Conversely, employees identified only local conditions, including economic, environmental, political, and technological factors, as well as critical incidents like higher-priority events or issues, as potential obstacles to implementation within the organization’s external context.

**Figure 3.**
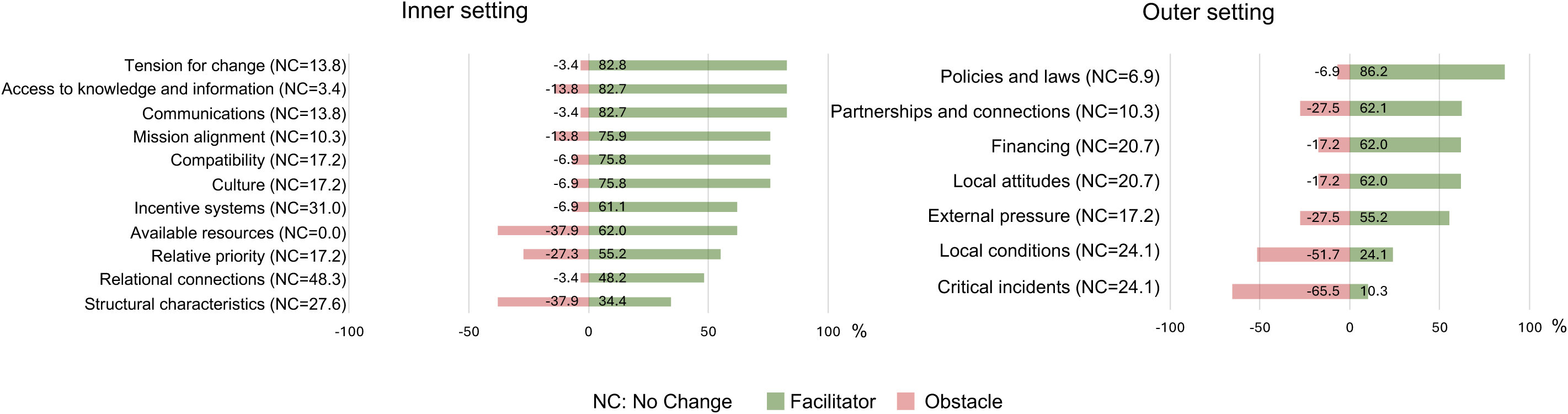
Percentage of employees perceiving contextual factors as facilitators or obstacles to implementation.

Figure 4 shows the effects reported by employees after participating in the three workshops and viewing the short videos. The effect most often reported by employees is increased awareness (79%). Greater awareness means that employees feel more involved in universal accessibility issues, and better understand their responsibilities in this area. Another effect is that employees say they have more information about universal accessibility (66%), implying an improvement in their knowledge of universal accessibility and the resources available to help them. In addition, employees still feel the need to better understand how to get personally involved (28%) and improve their practices (41%), implying a need for more tools from their work context, such as opportunities, additional training or personalized support.

**Figure 4.**
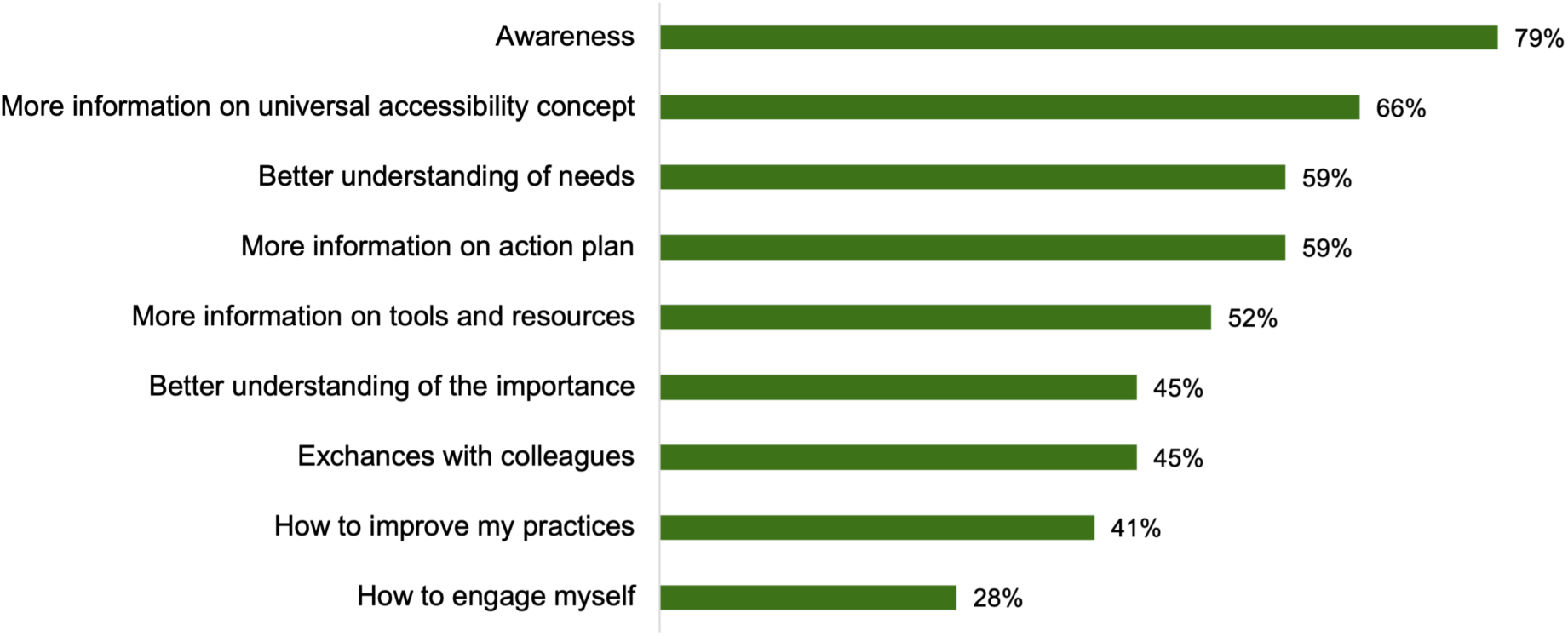
Percentage of employees seeing a change induced by tool implementation.

#### 2.2. Interviews

A total of 9 employees (n=3 from each team unit) participated in the interviews, that lasted in average 46 minutes, ranging from 28 to 75 minutes. The sample was composed of 7 women and 2 men. Two main themes emerge from the analysis: the format of the participatory workshops and the content of the short videos presented as knowledge mobilization tools.

##### Format of workshops

Employees gave positive feedback on the workshop format, finding the duration suitable. While virtual workshops were convenient, they often lacked interactive discussion compared to in-person sessions. Participants preferred smaller groups for richer discussions and increased participation. Opinions varied on the workshop frequency, with some preferring separate sessions for each video for ongoing discussion, while others favored a single longer session for deeper immersion. Consensus was reached on maintaining a week interval between sessions to aid information retention. Flexibility in adapting to team needs for broader implementation was deemed essential. Overall, participants found both the video content and workshop format beneficial for understanding universal accessibility’s relevance to their work and organization, suggesting improvements like involving more diverse perspectives and scheduling standalone workshops.

##### Content of short videos

Employees found that the short videos content positively impacted their understanding, beliefs, and awareness of universal accessibility and their ability to incorporate it into their work. Initially, the videos served as clear reminders of universal accessibility principles, enhancing participants’ comprehension, particularly with the provided examples. While some were already familiar with the concept, they appreciated the concise reinforcement. The videos also informed them about City initiatives and colleagues’ actions, illustrating varied implementation contexts and municipal-level concerns. In this regard, one participant stated, "*We can all contribute in our own way, after all, it’s great to know that!"*.

Participants recognized their role in contributing to universal accessibility and found understanding the role of accessibility managers beneficial, though they cautioned against overreliance. Additionally, the videos established a shared knowledge base among employees, facilitating resource sharing and preventing expertise loss. They also broadened awareness beyond wheelchair users, emphasizing universal accessibility’s relevance to diverse realities. One participant pointed out: *"It’s not about serving a minority; it’s about adapting for the greatest number of people who have many different realities".* Personal connections deepened participants’ understanding, highlighting its significance in both professional and personal spheres. Witnessing colleagues’ involvement in the videos helped participants envision integration into their own work and prioritize accordingly. However, they identified a need for additional tools to support integration efforts in the future. This made them aware of the breadth and scope of universal accessibility and helped them realize that it concerns everyone: *"Everyone benefits from [universal accessibility]; it touches everyone at the deepest level".* Moreover, several participants made connections with situations in their personal lives during the interviews, whether related to an aging relative or experiences they had encountered. One participant stated, *"I realized that everyone is directly or indirectly concerned, whether at work or in their personal life… universal accessibility allows for better representation of the reality of more people."*

## DISCUSSION

This study aimed to describe an adapted implementation strategy as carried out and to explore induced changes on municipal employees’ perceptions towards implementation of universal accessibility measures.

First, the results suggest that the short videos had positives effects. It enabled participants to broaden their understanding of universal accessibility, they became more aware of the issues and various achievements carried out by the municipal organization, they could establish a common knowledge, and it clarified their role and responsibility towards universal accessibility and how they could take action. These positive effects can be explained by the choice of knowledge mobilization tools and the implementation strategy. Indeed, the literature reports that the combined use of participatory workshops and postcard reminders to develop an overall implementation strategy is more effective to the adoption, implementation, and sustainability of an intervention [36–40]. It has also been reported that interactive strategies, such as the presentation of the tools followed by exchanges between colleagues in a workshop, are more effective than passive strategies for changing behavior [41, 42]. However, the positive results documented in this study could also be attributed to the co-design methodology employed for developing knowledge mobilization tools and the implementation strategy. While the engagement of stakeholders facilitated the implementation process, it is important to acknowledge the possibility of a social desirability bias influencing employee responses during the questionnaire and interviews. Additionally, teams who participated in the workshops and evaluation process may be used to this type of evaluation-consultation approach due to their extensive engagement with citizens. This familiarity could potentially influence results positively by enhancing their honesty and critical thinking, or negatively due to fatigue or a sense of task completion. While the selection of the evaluation methods can have impacted the findings, it did enhance feasibility within the partnership’s context and timeframe. Future research could benefit from stricter adherence to implementation evaluation guidelines, such as TiDiER [43] or STARI [44], to ensure more rigorous documentation of results and mitigate bias effectively.

Secondly, Nilsen and Bernhardsson [45] have demonstrated the importance of taking context into account to properly implement an intervention. Our study, carried out in a specific municipal organization, and considering its particular and complex context by involving the stakeholders in all stages, enabled us to develop a flexible implementation strategy, adapted and in line with the needs of the context in which our intervention was inserted. As mentioned in studies, implementation strategies that are based on and aligned with determinant assessment (barriers and facilitators), that engage stakeholders, such as in a steering committee, and that are feasible and evidence-based are shown to be more effective [36, 46]. The use of determinant models such as CFIR has enabled us to identify the barriers to be overcome, as well as the facilitators to be mobilized in the creation of knowledge mobilization tools and the implementation strategy to be adopted, as reported in other studies [47–49]. However, given the vast and changing context, the picture painted is a global one at the time the data were collected. Thus, it is important to pay attention to changes in organizational, political and team priorities in order to adapt strategy and tools over time.

Thirdly, this study draws a lesson from research carried out in partnership with municipal actors. As mentioned by Phillipson and colleagues [46], stakeholders engagement is perceived as bringing benefits to the process of knowledge exchange and production. Indeed, there are significant advantages to involving stakeholders in the development of implementation strategies, such as guaranteeing the feasibility, relevance, and acceptability of the strategies [36, 46]. The results of this study demonstrate these advantages through the creation of the steering committee and its significant involvement in this project. Latulippe and colleagues [25] also shed light on the best practices to be developed for beneficial partnership research. They mentioned it is essential to devote time to implement the project’s results in local environments, and to ensure that they are useful to the partners. The implementation strategy created to deliver the knowledge mobilization tools, as well as the pre-test in three administrative units, enabled us to ensure, with the partner, that the strategy was appropriate, adapted and matched their needs. Although partnership research offers numerous advantages, it also presents challenges such as communication difficulties, collaboration barriers, and differing perspectives that can lead to misunderstandings [25]. Therefore, establishing clear common objectives, methods of dissemination, defined roles, and transparent communication from the outset are essential for fostering a positive and impactful partnership. Despite these challenges, this study emphasizes the significant value of partnership research, especially with municipal organizations, in developing tools and assessing their impact faithfully and contextually to promote effective changes in practices.

Although this study shows positive results, there are some important limitations to consider for future research. First, the study’s focus on a specific municipal context may limit the generalizability of its findings to other organizational contexts with different levels of interest. The literature points out that it can be complicated to generalize an implementation in a given context, and that this requires a nuanced understanding of the dynamic and complex nature of the context [50, 51]. Also, given that the implementation pre-test took place within three administrative units already involved in universal accessibility initiatives, results showed that participants probably already had knowledge or skills in this field. These units were chosen to facilitate the testing of the animation guide developed by the members of the steering committee, who also held universal accessibility management positions within their unit. So, as the partners continue to roll out the implementation strategy of the knowledge mobilization tools developed, it’s possible that the results vary, particularly in terms of knowledge and skills, depending on the reality and commitment to universal accessibility of other administrative units. In fact, implementation strategies often work in a given context, according to certain mechanisms, and for a particular audience [52]. Finally, although the steering committee decided to use a logic model to structure the means and objectives of the implementation strategy, no evaluation framework was used to conduct the evaluation. For example, the RE-AIM framework is an evaluation tool that assesses reach, effectiveness, adoption, implementation, and maintenance and can be used in conjunction with the CFIR [53]. In the context of a partnership-based research, it was complicated for partners to rely on a strict evaluation framework, which is why the logic model was prioritized. However, the use of an evaluation framework could have enhanced the rigor of the evaluation process.

## CONCLUSION

The results of this study demonstrate that the objectives were achieved, given the positive feedback on the implementation strategy and the positive effects that the knowledge mobilization tools had on understanding, awareness, knowledge, and the sense of responsibility among municipal employees.

Maintaining adaptability within the implementation strategy emerges as pivotal, especially in accommodating variations across administrative units, such as their predominant engagement in fieldwork or online activities. It is anticipated that this flexibility inherent in the implementation approach will foster deeper engagement and a sense of ownership among team members. This, in turn, is poised to foster a more comprehensive participation and heightened sense of involvement among employees within each team.

## Data Availability

All data produced in the present work are contained in the manuscript

## ETHICAL APPROVAL AND CONSENT TO PARTICIPATE

This study was approved by the Comité d’Éthique de la recherche en réadaptation et intégration sociale of the Centre intégré universitaire de santé et services sociaux de la capitale-nationale (CER-S of CIUSSS-CN). The approval number is 2022-2507.

## CONSENT FOR PUBLICATION

Not applicable

## AVAILABILITY OF DATA AND MATERIALS

Not applicable

## AUTHORS’ CONTRIBUTIONS

MC conceptualized the study and was the lead author of the manuscript. FR and MEL contributed to methodology development, protocol development, and manuscript writing. All authors read and approved the final manuscript.

## ACKNOWLEGMENT

Not applicable

## REFERENCES

1. United Nations, *Convention on the Rights of Persons with Disabilities*, U. Nations, Editor. 2006: New York.

2. Eckhardt, J., C. Kaletka, and B. Pelka, Monitoring inclusive urban development alongside a human rights approach on participation opportunities. European Planning Studies, 2020. 28(5): p. 991–1009.

3. Afacan, Y. and S.O. Afacan, Rethinking social inclusivity: design strategies for cities. Proceedings of the Institution of Civil Engineers-Urban Design and Planning, 2011. 164(2): p. 93–105.

4. da Silva Pereira, R.S., et al., Municipalities and the promotion of architectural accessibility. Revista de Enfermagem Referência, 2018. 4(18): p. 29–38.

5. United Nations. *68% of the world population projected to live in urban areas by 2050, says UN*. Department of Economic and Social Affairs of United Nations 2018; Available from: https://www.un.org/development/desa/en/news/population/2018-revision-of-world-urbanization-prospects.html.

6. Frías-López, E. and J. Queipo-de-Llano, Methodology for ‘reasonable adjustment’ characterisation in small establishments to meet accessibility requirements: A challenge for active ageing and inclusive cities. Case study of Madrid. Cities, 2020. 103.

7. Itair, M., I. Shahrour, and I. Hijazi, The Use of the Smart Technology for Creating an Inclusive Urban Public Space. Smart Cities, 2023. 6(5): p. 2484–2498.

8. Baudot, P.Y., C. Borelle, and A. Revillard, Politiques du handicap Introduction. Terrains et Travaux : Revue de Sciences Sociales, 2013(23): p. 5–15.

9. Larkin, H., et al., Working with Policy and Regulatory Factors to Implement Universal Design in the Built Environment: The Australian Experience. Int J Environ Res Public Health, 2015. 12(7): p. 8157–71.

10. World Health Organization, Global Age-friendly Cities: A guide. 2007.

11. Yarskaya-Smirnova, V.N. and E.R. Yarskaia-Smirnova, THE RIGHT TO THE CITY IN THE PARADIGM OF MOBILITY. Vestnik Tomskogo Gosudarstvennogo Universiteta-Filosofiya-Sotsiologiya-Politologiya-Tomsk State University Journal of Philosophy Sociology and Political Science, 2018. 45: p. 165–173.

12. Rachele, J.N., et al., Feasibility and the care-full just city: Overlaps and contrasts in the views of people with disability and local government officers on social inclusion. Cities, 2020. 100.

13. Jackson, M.A., Accessing the Neighbourhood: Built Environment Performance for People with Disability. Architecture_MPS, 2019. 6(1).

14. Zallio, M. and P.J. Clarkson, Inclusion, diversity, equity and accessibility in the built environment: A study of architectural design practice. Building and Environment, 2021. 206: p. 108352.

15. Labbé, D., et al., Examining the Impact of Knowledge Mobilization Strategies to Inform Urban Stakeholders on Accessibility: A Mixed-Methods study. International journal of environmental research and public health, 2020. 17(5).

16. Franz. J., et al. Inclusive universal design practice and activism: a theoretical framework. in 3rd International Conference for Universal Design. 2010.

17. Aimar, D. and J.-F. Chanlat, La co-construction : une réponse à l’écart entre les discours et la réalité en matière de politique de handicap dans les organisations contemporaines. Question(s) de management, 2022. 38(1): p. 105–121.

18. Mosca, E.I. and S. Capolongo, *Universal Design-Based Framework to Assess Usability and Inclusion of Buildings*, in Computational Science and Its Applications – ICCSA 2020: 20th International Conference, Cagliari, Italy, July 1–4, 2020, Proceedings, Part V. 2020, Springer-Verlag: Cagliari, Italy. p. 316–331.

19. Haveri, A., Complexity in local government change. Public Management Review, 2006. 8(1): p. 31–46.

20. Walker, R.M., F.S. Berry, and C.N. Avellaneda, Limits on innovativeness in local government: examining capacity, complexity, and dynamism in organizational task environments. Public Administration, 2015. 93(3): p. 663–683.

21. Perkins, D.D. and M.A. Zimmerman, Empowerment theory, research, and application. American Journal of Community Psychology, 1995. 23(5): p. 569–579.

22. Ncoyini, S.S., ß and L. Cilliers, *Factors that influence knowledge management systems to improve knowledge transfer in local government: A case study of Buffalo City Metropolitan Municipality, Eastern Cape, South Africa*. SA Journal of Human Resource Management, 2020. 18(0).

23. Curl, A., J.D. Nelson, and J. Anable, Does Accessibility Planning address what matters? A review of current practice and practitioner perspectives. 2011. 2: p. 3–11.

24. Gamache, S., Routhier, F., Morales, E., Vandermissen, M-H., Leblond, J., Boucher, N., McFadyen, B.J., Noreau, L., Municipal Practices and needs regarding accessibility of pedestrian infrastructures for individuals with physical disabilities in Québec, Canada. Journal of Accessibility and Design for all, 2017. 4(1): p. 21–55.

25. Latulippe, K., et al., Facilitators and challenges in partnership research aimed at improving social inclusion of persons with disabilities. Disability and Rehabilitation, 2023: p. 1–12.

26. Wickremasinghe, D., et al., Taking knowledge users’ knowledge needs into account in health: an evidence synthesis framework. Health Policy and Planning, 2015. 31(4): p. 527–537.

27. Corcuff, M., et al., *Co-design knowledge mobilization tools for universal accessibility in municipalities.* Submitted to Frontiers in Rehabilitation Sciences, Section Disability, Rehabilitation, and Inclusion, 2024.

28. Hunter, A. and J.D. Brewer, Designing Multimethod Research, in The Oxford Handbook of Multimethod and Mixed Methods Research Inquiry, S.N. Hesse-Biber and R.B. Johnson, Editors. 2015, Oxford University Press. p. 185–205.

29. Kellogg Foundation, Logic Model Development Guide. 2004: W.K. Kellogg Foundation.

30. Lumivero. NVivo (Version 14). 2023.

31. Aalbers, M., A South African Municipality Mapping the Way Forward for Social Inclusion Through Universal Design. Studies in health technology and informatics, 2016. 229: p. 53–62.

32. Corcuff, M., et al., *Municipalities’ strategies to implement universal accessibility measures: A scoping review.* Canadian journal of urban research, Revue canadienne de recherche urbaine, 2023. 32(2).

33. Damschroder, L.J., et al., Conceptualizing outcomes for use with the Consolidated Framework for Implementation Research (CFIR): the CFIR Outcomes Addendum. Implementation Science, 2022. 17(1).

34. Lozano, L.M., E. García-Cueto, and J. Muñiz, Effect of the Number of Response Categories on the Reliability and Validity of Rating Scales. Methodology, 2008. 4(2): p. 73–79.

35. Krosnick, J.A. and S. Presser, *Question and Questionnaire Design*, in Handbook of Survey Research (2nd Edition), J.D.W.a.P.V. Marsden, Editor. 2009, Elsevier: San Diego, CA.

36. Proctor, E.K., B.J. Powell, and J.C. McMillen, Implementation strategies: recommendations for specifying and reporting. Implementation Science, 2013. 8(1): p. 139.

37. Grimshaw, J.M., et al., Effectiveness and efficiency of guideline dissemination and implementation strategies. Health Technol Assess, 2004. 8(6): p. iii–iv, 1-72.

38. Menon, A., et al., Strategies for rehabilitation professionals to move evidence-based knowledge into practice: a systematic review. J Rehabil Med, 2009. 41(13): p. 1024–32.

39. Bernhardsson, S., et al., Determinants of guideline use in primary care physical therapy: a cross-sectional survey of attitudes, knowledge, and behavior. Phys Ther, 2014. 94(3): p. 343–54.

40. Baker, R., et al., Tailored interventions to address determinants of practice. Cochrane Database Syst Rev, 2015. 2015(4): p. Cd005470.

41. Powell, B.J., et al., A refined compilation of implementation strategies: results from the Expert Recommendations for Implementing Change (ERIC) project. Implementation Science, 2015. 10(1): p. 21.

42. Prior, M., M. Guerin, and K. Grimmer-Somers, The effectiveness of clinical guideline implementation strategies--a synthesis of systematic review findings. J Eval Clin Pract, 2008. 14(5): p. 888–97.

43. Hoffmann, T.C., et al., Better reporting of interventions: template for intervention description and replication (TIDieR) checklist and guide. BMJ : British Medical Journal, 2014. 348: p. g1687.

44. Pinnock, H., et al., Standards for Reporting Implementation Studies (StaRI) Statement. BMJ, 2017. 356: p. i6795.

45. Nilsen, P. and S. Bernhardsson, Context matters in implementation science: a scoping review of determinant frameworks that describe contextual determinants for implementation outcomes. BMC Health Serv Res, 2019. 19(1): p. 189.

46. Phillipson, J., et al., Stakeholder engagement and knowledge exchange in environmental research. J Environ Manage, 2012. 95(1): p. 56–65.

47. Adamu, A.A., et al., Using the consolidated framework for implementation research (CFIR) to assess the implementation context of a quality improvement program to reduce missed opportunities for vaccination in Kano, Nigeria: a mixed methods study. Human Vaccines & Immunotherapeutics, 2020. 16(2): p. 465–475.

48. Breimaier, H.E., et al., The Consolidated Framework for Implementation Research (CFIR): a useful theoretical framework for guiding and evaluating a guideline implementation process in a hospital-based nursing practice. BMC Nursing, 2015. 14(1).

49. Keith, R.E., et al., Using the Consolidated Framework for Implementation Research (CFIR) to produce actionable findings: a rapid-cycle evaluation approach to improving implementation. Implementation Science, 2017. 12(1).

50. Mielke, J., et al., Understanding dynamic complexity in context—Enriching contextual analysis in implementation science from a constructivist perspective. Frontiers in Health Services, 2022. 2.

51. Pfadenhauer, L.M., Conceptualizing Context and Intervention as a System in Implementation Science: Learning From Complexity Theory Comment on "Stakeholder Perspectives of Attributes and Features of Context Relevant to Knowledge Translation in Health Settings: A Multi-countr. International Journal of Health Policy and Management, 2021.

52. Jagosh, J., et al., A realist evaluation of community-based participatory research: partnership synergy, trust building and related ripple effects. BMC Public Health, 2015. 15(1): p. 725.

53. King, D.K., et al., Planning for Implementation Success Using RE-AIM and CFIR Frameworks: A Qualitative Study. Frontiers in Public Health, 2020. 8.

